# Increasing trend in fusidic acid resistance among MRSA isolates in the Netherlands, 2016-2023

**DOI:** 10.1101/2025.09.12.25335629

**Authors:** F Velthuis, IM Nauta, W Altorf-van der Kuil, DW Notermans, RD Zwittink, AF Schoffelen, SC de Greeff, the Infectious Diseases Surveillance Information System-Antimicrobial Resistance (ISIS-AR) Study Group

## Abstract

**Objectives:** Recently, several methicillin-resistant *Staphylococcus aureus* (MRSA) community outbreaks occurred in the Netherlands, including one caused by an impetigo-causing MRSA strain resistant to fusidic acid. Since fusidic acid and flucloxacillin are the main treatment options for impetigo, increasing resistance limits treatment possibilities. We examined trends in fusidic acid resistance levels among MRSA isolates in the Netherlands.

**Methods:** Data on routine bacteriological cultures between 2016-2023 from 30 laboratories were extracted from the national surveillance system on antimicrobial resistance (ISIS-AR). Fusidic acid resistance rates per year were calculated both overall and per age group for all MRSA isolates, and more specific, for the subset of MRSA isolates from wound/pus/skin samples collected by general practitioners (WPS-GP-samples). Trends were determined using logistic regression and compared with trends among methicillin-susceptible *S. aureus* (MSSA) isolates.

**Results:** We found an increasing trend in fusidic acid resistance among MRSA isolates from 15% in 2016 to 30% in 2023 (p<0.001) which differed significantly from the trend among MSSA isolates (p<0.001). An increase was also found in MRSA WPS-GP-samples, both among young children and the population of 13-64 years old, but not among elderly. The trends remained significant after exclusion of isolates associated with known fusidic acid-resistant MRSA outbreaks, both among MRSA isolates overall (OR = 1.10, 95% CI: 1.07-1.14, p<0.001) and among MRSA WPS-GP-samples (OR = 1.14, 1.07-1.21, p<0.001).

**Conclusions:** In conclusion, an increasing trend in fusidic acid resistance was found among MRSA isolates. Since impaired treatment for impetigo might ease the spread of (fusidic acid-resistant) MRSA, extra vigilance is warranted.

## Introduction

*Staphylococcus aureus* is a commensal bacterium which can act as a pathogen causing skin infections like impetigo. ^1,2^ Topical fusidic acid is the first-line treatment of impetigo in the Netherlands, as well as in other European countries, ^3,4^ and is often prescribed empirically by general practitioners (GPs). ^5^ In the Netherlands, oral flucloxacillin is the second choice in case of treatment failure (only effective against methicillin-susceptible *S. aureus* (MSSA)).

Transmission of impetigo with fusidic acid-resistant *S. aureus* has been reported in the Netherlands and other European countries, ^6-8^ and a slight increase of fusidic acid resistance was found among wound and pus isolates collected by GPs between 2019 (20%) and 2023 (23%) in the Netherlands. ^9^ In Norway and Denmark, countries with similar levels of MRSA as the Netherlands, fusidic acid resistance was found on the rise between 2016 to 2023 among methicillin-resistant *S. aureus* (MRSA) isolates specifically. ^10-12^ Besides, multiple outbreaks with fusidic acid-resistant MRSA have been reported to the ECDC in recent years: amongst others an outbreak of impetigo cases in the Netherlands in 2019 caused by an MRSA MT4627 strain. ^13^ This raised concern as fusidic acid and flucloxacillin are both ineffective in this case. Especially the increase of fusidic acid resistance in MRSA is worrisome as this may lead to unnoticed spread of MRSA. Since GPs in the Netherlands generally only take cultures in case of treatment failure, patients with insufficiently treated impetigo will stay contagious for a longer period, thereby facilitating transmission of MRSA.

The National Institute for Public Health and the Environment (RIVM) carries out surveillance on antimicrobial resistance (AMR) in the Netherlands based on data from the Infectious Diseases Surveillance Information System – Antimicrobial Resistance (ISIS-AR). The aim of this study was to investigate fusidic acid resistance levels among MRSA isolates in the Netherlands between 2016-2023, using data from this surveillance system. These data were compared with fusidic acid resistance levels among MSSA isolates.

## Materials and methods

### Data collection

ISIS-AR systematically collects, integrates, and analyses microbiological and epidemiological data from medical microbiology laboratories (MMLs) across the country. The database includes detailed information from routine diagnostics of positive bacterial cultures, including bacterial species identification, antimicrobial susceptibility test results (AST), patient demographics (such as age, sex, and type of healthcare setting where the sample was taken), specimen type (e.g., blood, urine), and sampling date. ^14^ From the 48 MMLs in the Netherlands 37 continuously delivered data from 2016 until 2023. The first diagnostic (infection-related) *S. aureus* isolate per patient per year was selected as well as information on AST for fusidic acid, cefoxitin, flucloxacillin and oxacillin and results from confirmation tests for MRSA. To minimize bias, an antibiotic agent should be tested in at least 50% of the isolates and at least 80% of the crude test results should be reinterpretable by EUCAST breakpoints for each MML. This resulted in 30 MMLs eligible for inclusion in the analyses.

### Calculation of resistance levels

An *S. aureus* isolate was considered MRSA if the presence of a *mecA* or m*ecC* gene or pbp2 production was demonstrated, or in case data on such confirmation tests was unavailable, if resistance to cefoxitin based on laboratory interpretation was demonstrated, or if also unavailable, if resistance to flucloxacillin/oxacillin based on laboratory interpretation was demonstrated. Susceptibility to topical fusidic acid was estimated based on reinterpretation of raw testing values according to EUCAST version 14.0 (R>0.5 mg/L). Within both the MRSA and MSSA group the prevalence of fusidic acid resistance was calculated as percentage of the total number of isolates. Additionally, fusidic acid resistance percentages were calculated for MRSA isolates from wound/pus/skin samples collected by GPs (hereafter referred to as WPS-GP-samples). Lastly, WPS-GP-samples were categorized by age group (0-12, 13-64 and ≥65 years old) since resistance patterns and type of diagnosis might be different between age groups. We assume that the approximation of impetigo-related isolates is most accurate for the WPS-GP-samples within the youngest age group. Resistance was calculated per year.

### Calculation of time trends

Time trends were assessed using logistic regression models for both MSSA and MRSA overall and for WPS-GP isolates, overall and stratified by age group, covering the period from 2016 to 2023. Models were fitted only where the number of susceptible and resistant isolates for all strata for all years exceeded 10. Logistic regression was performed with the antimicrobial susceptibility test (AST) interpretation for topical fusidic acid as the binary outcome (resistant versus non-resistant) and the Akaike Information Criterion (AIC) was used to choose the best fitting model. In case a non-linear trend fitted the data best for one of the groups (i.e. MSSA or MRSA overall and WPS-GP isolates, overall or stratified by age group) both a linear and non-linear trend were calculated for all groups. The non-linear trends were calculated per two years for interpretation purposes. Each analysis was corrected for age and sex. For comparing trends between MRSA and MSSA isolates an interaction term for MRSA/MSSA was added to the model. Calculations were performed in R version 4.4.3. A two-sided p-value of <0.05 was considered statistically significant.

### Sensitivity analysis

To minimize the potential impact of overrepresentation of MRSA outbreak-related isolates with fusidic acid resistance on observed trends, we repeated the analyses after excluding these outbreak-related isolates. In the Netherlands, multiple-locus variable number of tandem repeat analysis (MLVA)-complex MC0005 and MLVA-type MT4627 (part of MLVA-complex MC0030) are associated with fusidic acid resistance (∼50% and 100% fusidic acid-resistant in 2023 respectively) and related to community MRSA outbreaks. ^13^ To identify isolates in the ISIS-AR database belonging to this complex and type, molecular typing results from the Dutch national MRSA surveillance system (Type-Ned) were linked to the ISIS-AR dataset. ^15^ First, isolates were matched based on laboratory, sample number, isolate number and species. If matching failed, isolates were matched based on patient characteristics.

## Results

The number of isolates included and associated patient characteristics are shown in Table 1. Overall we included 344,387 MSSA isolates from 308,606 different patient, 7,553 MRSA isolates from 7,150 patients, 59,498 MSSA-WPS-GP isolates from 57,511 patients and 1,677 MRSA-WPS-GP isolates from 1,648 patients. Of the MRSA-WPS-GP isolates 242 (14.4%) came from patients aged 0-12 years, 1,168 (69.6%) from patients aged 13-64 years and 267 (15.9%) from patients aged ≥ 65 years.

**Table 1.** Numbers of MSSA and MRSA isolates overall and from wound/pus/skin samples collected by GPs (WPS-GP), and related patient characteristics.

|  | MSSA |  | MRSA |  | MSSA-WPS-GP |  | MRSA-WPS-GP |  |
| --- | --- | --- | --- | --- | --- | --- | --- | --- |
|  | N | % | N | % | N | % | N | % |
| **Patients** | 308,606 | - | 7,150 | 2.3* | 57,511 | - | 1,648 | 2.8* |
| **Isolates** | 344,387 | - | 7,553 | 2.1* | 59,498 | - | 1,677 | 2.7* |
| **Age** |  |  |  |  |  |  |  |  |
| 0-12 years | 30,068 | 8.7 | 751 | 9.9 | 10,254 | 17.2 | 242 | 14.4 |
| 13-64 years | 164,154 | 47.7 | 4,470 | 59.2 | 31,788 | 53.4 | 1,168 | 69.6 |
| ≥65 years | 150,165 | 43.6 | 2,332 | 30.9 | 17,456 | 29.3 | 267 | 15.9 |
| **Sex** |  |  |  |  |  |  |  |  |
| Male | 182,536 | 53.0 | 4,485 | 59.4 | 28,596 | 48.1 | 972 | 58.0 |
| Female | 161,851 | 47.0 | 3,068 | 40.6 | 30,902 | 51.9 | 705 | 42.0 |
| **Sample material** |  |  |  |  |  |  |  |  |
| Blood | 16,032 | 4.7 | 234 | 3.1 | - | - | - | - |
| Respiratory | 49,907 | 14.5 | 1,152 | 15.3 | - | - | - | - |
| Wound/pus/skin | 214,086 | 62.2 | 4,816 | 63.8 | 59,498 | 100 | 1,677 | 100 |
| Other | 64,362 | 18.7 | 1,351 | 17.9 | - | - | - | - |
| **Healthcare setting** |  |  |  |  |  |  |  |  |
| General practitioner | 98,369 | 28.6 | 2,477 | 32.8 | 59,498 | 100 | 1,677 | 100 |
| Inpatient department | 61,417 | 17.8 | 1,360 | 18.0 | - | - | - | - |
| Outpatient department | 142,525 | 41.4 | 2,655 | 35.2 | - | - | - | - |
| Other | 42,076 | 12.2 | 1,061 | 14.0 | - | - | - | - |

\*Percentage MRSA among the number of patients and the number of isolates, overall and for the WPS-GP selection group.

The annual fusidic acid resistance percentage increased significantly from 15% in 2016 to 30% in 2023 among MRSA isolates (OR = 1.13, 95% CI: 1.09-1.16, p<0.001) and from 10% in 2016 to 12% in 2023 among MSSA isolates (OR = 1.01, 95% CI: 1.00-1.02, p<0.001) as determined by the linear regression model (Figure 1A). The difference between the trends of MRSA and MSSA was statistically significant (OR = 1.11, 95% CI: 1.08-1.14, p<0.001). A rising trend was also found among WPS-GP-samples and was more profound among MRSA isolates (OR = 1.16, 95% CI: 1.11-1.23, p<0.001) compared to MSSA isolates (OR = 1.02, 95% CI: 1.01-1.03, p<0.001) (Figure 1B). The difference between trends between both groups was again statistically significant (OR = 1.14, 95% CI: 1.08-1.20, p<0.001). The quadratic model best fitted the time trend for both the overall MRSA and MSSA isolates when comparing the AIC with the linear model. Therefore, additional ORs were calculated per two years which were highest and significant for 2022-2023 vs 2016-2017 (Supplementary Table 1).

**Figure 1.**
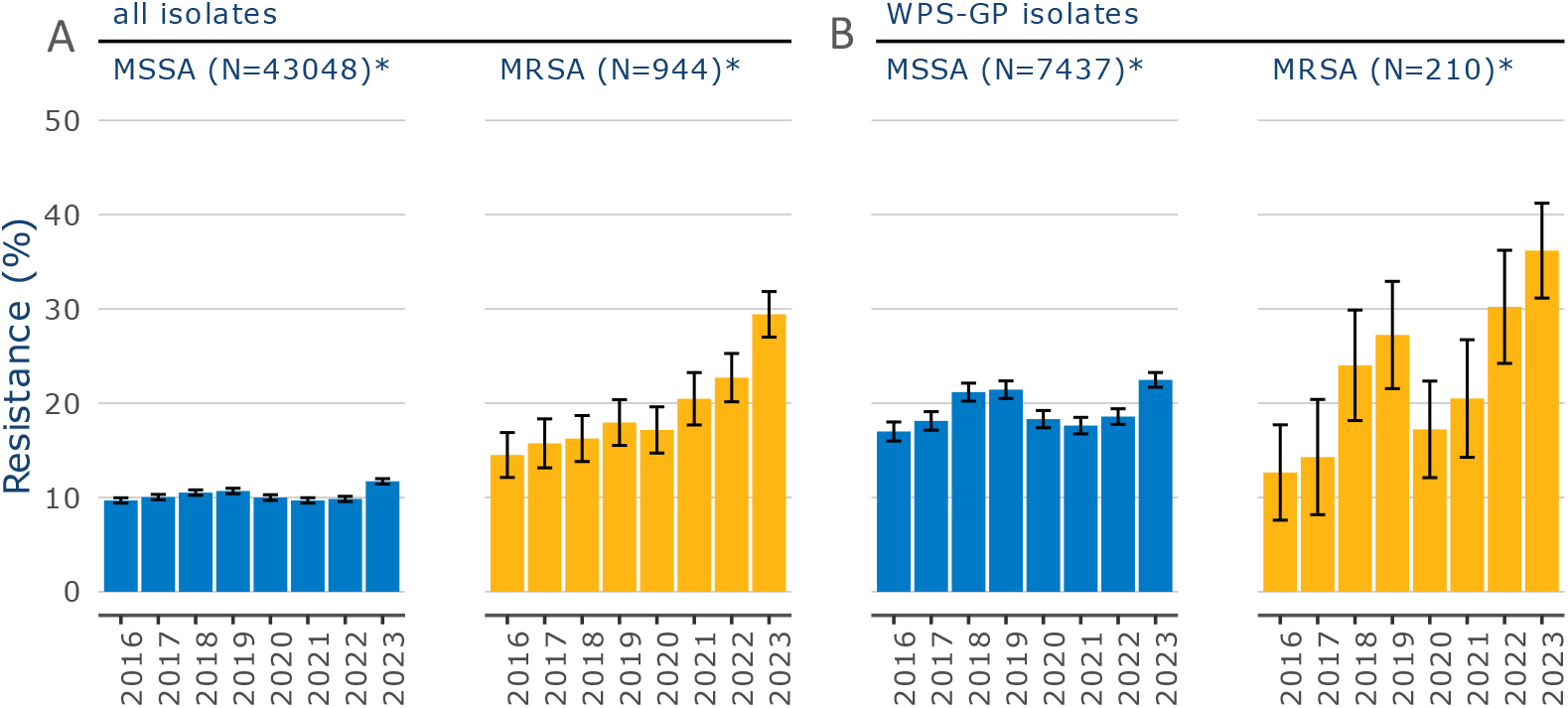
Fusidic acid resistance percentages in methicillin-susceptible S. aureus (MSSA) and methicillin-resistant S. aureus (MRSA) isolates between 2016 to 2023 in the Netherlands. Left two panels (A) show isolates from all materials and healthcare settings (all isolates). Right two panels (B) show isolates from wound/pus/skin samples collected by the general practitioner (WPS-GP isolates). The error bars show the 95% confidence intervals. *Mean number isolates tested for fusidic acid per year.

After stratification of WPS-GP-samples by age group, different resistance trends over time could be observed per age group (Figure 2). As the number of resistant isolates was below 10 for multiple years, no trend analyses were performed for the separate age groups. Visually, fusidic acid resistance increased over time among 0-12 year olds, with a peak in 2019, while among 13-64 year olds no such peak was visible and the trend of resistance level seemed less steep. Resistance levels for the group ≥65 year old remained more stable.

**Figure 2.**
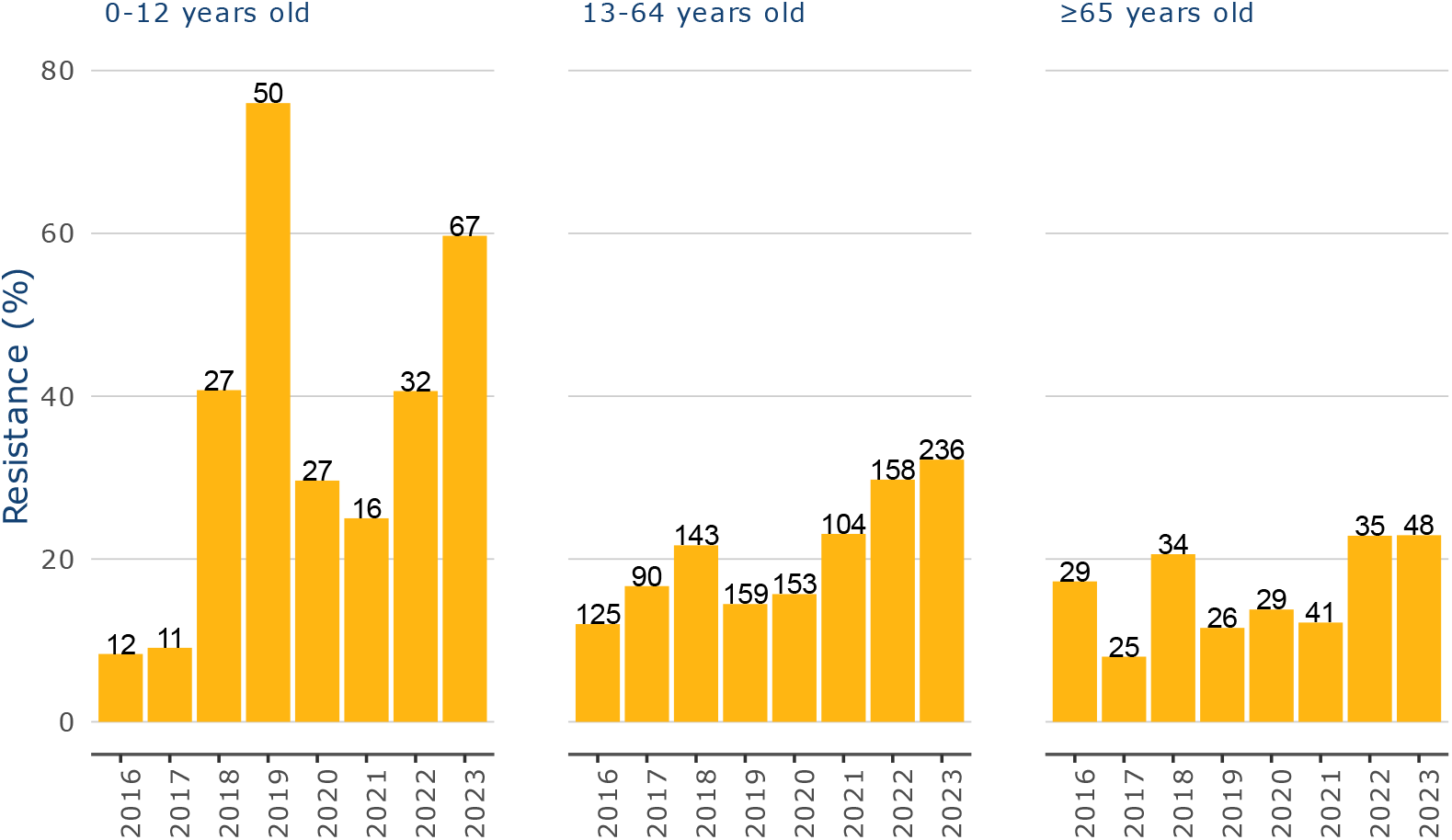
Fusidic acid resistance rates in methicillin-resistant S. aureus (MRSA) wound/pus/skin samples collected by the general practitioner between 2016 to 2023 per age category in the Netherlands. Age categories are defined as follows: 0-12 years old, 13-64 years old and ≥65 years old. Numbers above bars show the total number of MRSA isolates tested for fusidic acid susceptibility per year.

### Sensitivity analysis

Of the patients with MRSA and MRSA-WPS-GP, 5,737 (80.2%) and 1,384 (84.0%) could be linked to an MRSA isolate in Type-Ned with molecular typing data, respectively. The distribution of age, sex, sample material and healthcare setting were similar between the full MRSA dataset and the MRSA dataset with linked molecular typing data, as well as between the MRSA-WPS-GP dataset and the MRSA-WPS-GP with linked molecular typing data (data not shown).

When excluding MLVA-complex MC0005 and MLVA-type MT4627 (part of MC0030) from the overall MRSA dataset, we found a less steep but still significant trend for fusidic acid resistance percentages. The percentage increased from 12% in 2016 to 22% in 2023 (OR = 1.10, 95% CI: 1.07-1.14, p<0.001) (Figure 3). When excluding MLVA-complex MC0005 and MLVA-type MT4627 from the MRSA-WPS-GP dataset we again found a less steep but still significant trend with fusidic acid resistance percentages increasing from 9% in 2016 to 21% in 2023 (OR = 1.14, 95% CI: 1.07-1.21, p<0.001). The proportions of MLVA-complexes over time for both the overall MRSA selection group and the WPS-GP selection group are shown in Supplemental Figure 1 and 2 for fusidic acid-resistant MRSA and Supplemental Figure 3 and 4 for fusidic acid-resistant and -susceptible MRSA combined.

**Figure 3.**
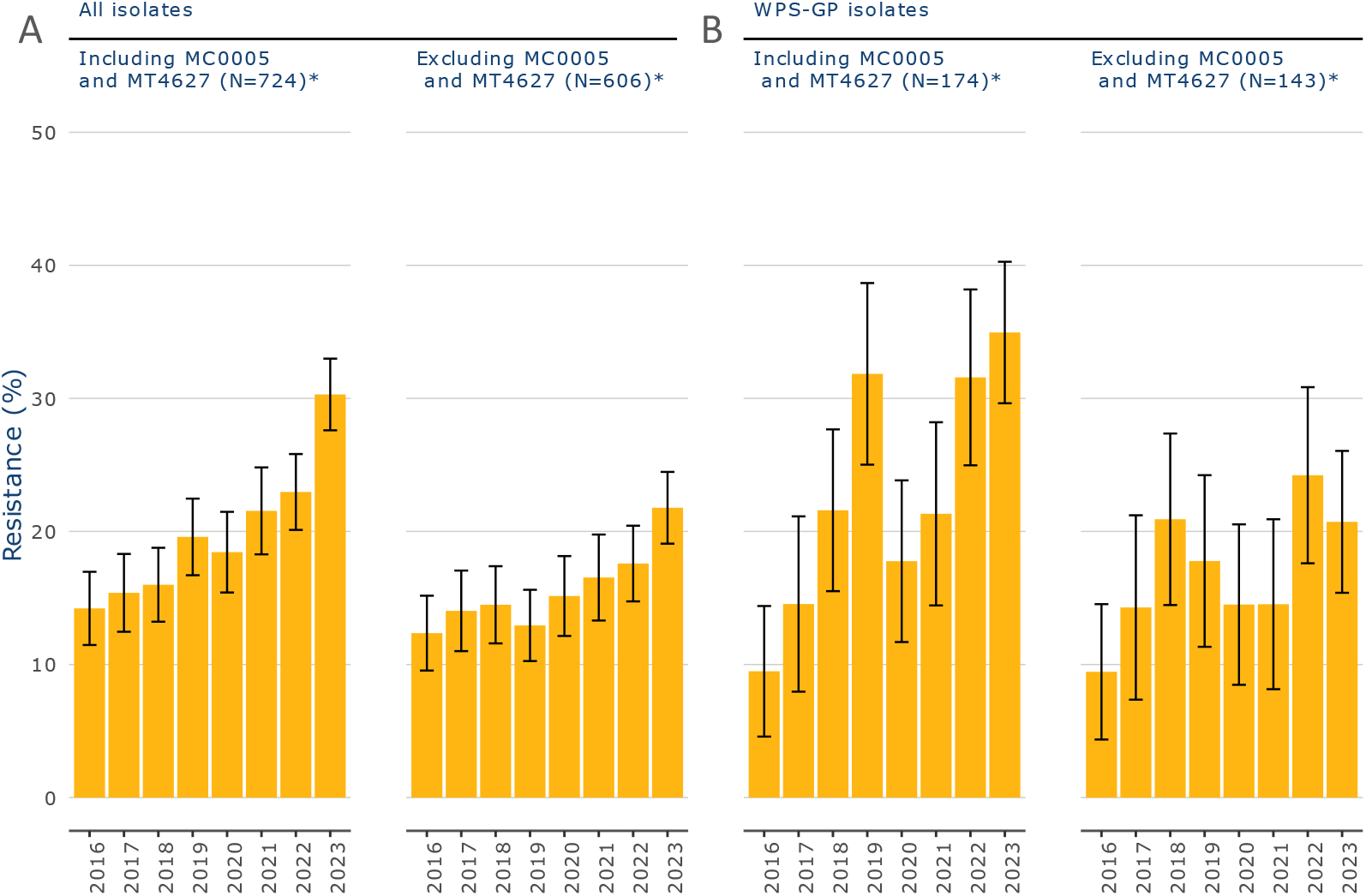
Fusidic acid resistance percentages in methicillin-resistant S. aureus (MRSA) linked to Type-Ned between 2016 and 2023 in the Netherlands. Left two panels (A) show isolates from all materials and healthcare settings (all isolates) including and excluding MC0005 and MT4627. Right two panels (B) show isolates from wound/pus/skin samples collected by the general practitioner (WPS-GP isolates) including and excluding MC0005 and MT4627. The error bars show the 95% confidence intervals. *Mean number isolates tested for fusidic acid per year.

## Discussion

The aim of this study was to investigate trends in fusidic acid resistance levels among MRSA isolates over a period of eight years in the Netherlands using surveillance data from ISIS-AR. The results show an increase in fusidic acid resistance among MRSA isolates from 15% to 30%, with the increase being more pronounced compared to fusidic acid resistance in MSSA. Analyses focusing on wound/pus/skin isolates from GP patients showed similar findings, with a sudden increase from 2018 in young children (≤12 years old). In sensitivity analyses, upon exclusion of MLVA-complex MC0005 and MLVA-type MT4627 which are associated with fusidic acid-resistant MRSA outbreaks, the rising trend was still visible, although somewhat less steep. This finding suggests that the observed increase in resistance could not be fully attributed to this MLVA-complex and MLVA-type.

In the Netherlands, fusidic acid is the first choice treatment for impetigo. In case of treatment failure, flucloxacillin is prescribed. However, if the impetigo is caused by MRSA, treatment with flucloxacillin will also not be effective. The more pronounced increase in fusidic acid resistance among MRSA isolates compared to MSSA isolates may be explained by the ineffective treatment of impetigo infections caused by fusidic acid-resistant MRSA. Because patients with fusidic acid-resistant MRSA impetigo cannot be treated with either the first- or second-line therapies, they are likely to remain contagious within the community for a longer period, facilitating transmission and contributing to an increased proportion of resistant isolates. In contrast, impetigo caused by MSSA can be successfully treated with flucloxacillin if the first-line therapy fails, thereby reducing the likelihood of ongoing transmission. Even though infections with MRSA are less frequent than MSSA, this poses a potential threat to public health as this eases spread of MRSA.

The increase in fusidic acid resistance among MRSA isolates is also seen in other European countries, for example in Denmark (from 18% to 26% between 2016-2023) and Norway (from 12% to 16% between 2016-2023). ^10-12^ Meanwhile, the percentage fusidic acid resistance among *S. aureus* bacteraemia cases in Denmark remained stable between 2014 and 2023 at 12% which corresponds to the resistance percentage that we found among MSSA isolates, although we did not analyse trends for bacteraemia cases specifically. ^11^ Similarly, a stable trend was found in Norway in fusidic acid resistance among *S. aureus* wound isolates between 2016 and 2023 around 7%. ^10,12^ Together, these results support our finding that the trend of fusidic acid resistance among MRSA isolates increases and can differ from MSSA isolates.

The steep increases in resistance visible in young children (age 0-12 years old) in 2018 with a peak around 2019 and 2023 may imply transmission of a fusidic acid-resistant MRSA strain. The peak in 2019 is likely influenced by the outbreak in the Netherlands that year. ^13^ Whereas impetigo is easily transmitted between children and the increased spread of fusidic acid-resistant MRSA may somewhat be expected, the rising resistance trend among 13-64 years old is more remarkable. This suggests that there may be a general increase in fusidic acid resistance among MRSA isolates that cannot be directly attributed to the known outbreak cluster. A possible explanation might be that the outbreak in 2019 led to an increased spread of fusidic acid-resistant MRSA causing other wound/pus/skin infections next to impetigo.

The increase of fusidic acid resistance among MRSA isolates restricts the number of treatment options for impetigo. Possible alternative topical antibiotics are mupirocin and retapamulin. In the Netherlands, however, mupirocin is primarily used for eradication of MRSA carriage in healthcare professionals (also known as ‘the search and destroy’ policy). ^16^ Due to this policy, MRSA is still present at very low levels in the Netherlands. ^17^ Only in recurrent cases of impetigo with *S. aureus*, mupirocin is advised. In France, mupirocin is the first choice treatment for impetigo infections due to already high levels of fusidic acid resistance. ^18^ Retapamulin has been shown to be as effective as fusidic acid in clearing an impetigo infection in a randomized non-inferiority study across multiple countries. ^19^ However, data on its effectiveness in eradicating MRSA impetigo infections are limited. As a reaction to the rise in fusidic acid resistance, a single blind randomised controlled trial was set up in New Zealand. ^20^ The aim of this RCT was to determine non-inferiority treatment of antiseptics like hydrogen peroxide since this type of treatment could lower the risk of acquiring resistance. The results of this study are still awaited. Overall, topical treatment options for MRSA impetigo infections are limited as mupirocin is advised to be used only in specific cases and more research is needed to prove efficacy of alternative topical treatments.

One of the strengths of this study is the use of a large national database with a high coverage of MMLs across the country. ^14^ Incoming data are checked on a regular basis and verified in consultation with MMLs before validated and incorporated into the database. This quality check process results in a database in which inaccuracies are minimized. Another strength is that isolates from the ISIS-AR database were linked to data in the Type-Ned database, which enabled us to include molecular typing data in our analyses and exclude isolates associated with fusidic acid-resistant MRSA outbreaks.

A limitation of this study is the lack of clinical data within ISIS-AR. By selecting WPS-GP-samples we attempted to approximate the population with impetigo as closely as possible. However, part of the selection might be non-impetigo related, especially among the ≥65 age group. In this age group we also expect patients with other infections such as surgical site infections or diabetic foot ulcers ulcers. Another limitation is that the protocol for the national Type-Ned MRSA surveillance prescribes to send only one MRSA isolate per patient per three years for molecular typing. In addition to this, ISIS-AR collects the first *S. aureus* isolate per patient per year, either MSSA or MRSA. Therefore, only part of the ISIS-AR dataset could be linked to the Type-Ned dataset and only for a part of all MRSA isolates from ISIS-AR molecular data was available. However, patient characteristics from the linked sub selection were similar to the selection for the main analyses, meaning that differences in outcomes cannot be attributed to different patient populations.

For the interpretation of the resistance percentages it should be considered that GPs usually do not take cultures for impetigo, unless after treatment failure, resulting in overestimation of resistance percentages in WPS-GP-samples. We assumed that culture practice did not change over time, except during the outbreak in 2019 in that specific geographical area, ^13^ and that selective sampling would not have influenced trends over time. It is however possible that the outbreak led to more awareness of fusidic acid-resistant MRSA among GPs, resulting in a sustained, low-threshold approach to performing cultures.

To conclude, this study shows an increasing trend in fusidic acid resistance among MRSA isolates, which is more profound compared to the trend in MSSA isolates and cannot be fully explained by a specific MLVA-complex or -type associated with fusidic acid-resistant MRSA outbreaks. Although MRSA levels in the Netherlands are still low, the increase of fusidic acid resistance among MRSA isolates might pose a public health threat as impaired treatment can lead to the spread of MRSA. Based on these findings, surveillance to timely detect fusidic acid resistance among *S. aureus* isolates is warranted to optimize treatment guidelines. Furthermore, fusidic acid resistance should be monitored for MRSA and MSSA separately, as this study shows that resistance levels differ. When focusing on *S. aureus* isolates overall, this steeper increase in MRSA would not have been noticed. It may be advisable to culture skin lesions caused by impetigo at an earlier stage, than only after treatment failure. As a reaction to the outcomes of this study, the Dutch General Practitioner Federation indeed advised GPs to culture patients with impetigo infection prior to antibiotic treatment in case of increased risk of MRSA or in case the patient may be part of a regional cluster of patients with fusidic acid treatment failure. ^21^ Future research could aim to acquire an unbiased selection of samples. For example by sampling all patients with impetigo at GPs. This would gain insight into the actual resistance rates and the impact of impetigo outbreaks on these rates.

## Data Availability

All data produced in the present study are available upon reasonable request to the authors

## Acknowledgements

We would like to take this opportunity to thank all contributing MMLs for providing data to ISIS-AR. The findings of this study were previously presented as a poster at ESCMID Global 2025.

## Transparency declarations

### Members of the ISIS-AR study group

J.W.T. Cohen Stuart, Noordwest Ziekenhuisgroep, Department of Medical Microbiology, Alkmaar

D.C. Melles, Meander Medical Center, Department of Medical Microbiology, Amersfoort

K. van Dijk, Amsterdam UMC, University of Amsterdam, Department of Medical Microbiology and Infection Prevention, Amsterdam Infection and Immunity Institute, Amsterdam

R. Schade, Amsterdam UMC, University of Amsterdam, Department of Medical Microbiology and Infection Prevention, Amsterdam Infection and Immunity Institute, Amsterdam

A. Alzubaidy, Atalmedial, Department of Medical Microbiology, Amsterdam

M. Scholing, OLVG Lab BV, Department of Medical Microbiology, Amsterdam

S.D. Kuil, Public Health Service, Public Health Laboratory, Amsterdam

G.J. Blaauw, Gelre Hospitals, Department of Medical Microbiology and Infection prevention, Apeldoorn

ISIS-AR Projectteam: W. Altorf - van der Kuil, S.M. Bierman, S.C. de Greeff, S.R. Groenendijk, R. Hertroys, L. Kruithof, I.M. Nauta, D.W. Notermans, J. Polman, W.J. van den Reek, A.F. Schoffelen, F. Velthuis, C.C.H. Wielders, B.J. de Wit, R.E. Zoetigheid, Centre for Infectious Disease Control (CIb), National Institute for Public Health and the Environment (RIVM), Bilthoven, The Netherlands

W. van den Bijllaardt, Microvida Amphia, Laboratory for Microbiology and Infection Control, Breda

E.M. Kraan, IJsselland hospital, Department of Medical Microbiology, Capelle a/d IJssel

M.B. Haeseker, Reinier de Graaf Group, Department of Medical Microbiology, Delft

J.M. da Silva, Deventer Hospital, Department of Medical Microbiology, Deventer

E. de Jong, Slingeland Hospital, Department of Medical Microbiology, Doetinchem

B. Maraha, Albert Schweitzer Hospital, Department of Medical Microbiology, Dordrecht

M.P.A. van Meer, Department of Medical Microbiology and Immunology, Dicoon, Elst

B.B. Wintermans, Admiraal De Ruyter Hospital, Department of Medical Microbiology, Goes

V. Hira, Groene Hart Ziekenhuis, Department of Medical Microbiology and Infection Prevention, Gouda

A.E. Muller, Haaglanden MC, Department of Medical Microbiology, ‘s-Gravenhage

M. Wong, Haga Hospital, Department of Medical Microbiology, ‘s-Gravenhage

P. Huizinga, Certe, Groningen

E. Bathoorn, University of Groningen, University Medical Center, Department of Medical Microbiology, Groningen

M. Lokate, University of Groningen, University Medical Center, Department of Medical Microbiology, Groningen

J. Sinnige, Regional Public Health Laboratory Haarlem, Haarlem

L.E.A. Bank, St Jansdal Hospital, Department of Medical Microbiology, Harderwijk

F.W. Sebens, Labmicta, Hengelo

E. Kolwijck, Jeroen Bosch Hospital, Department of Medical Microbiology and Infection Control, ‘s-Hertogenbosch

E.A. Reuland, CBSL, Tergooi MC, Department of Medical Microbiology, Hilversum

J.W. Dorigo-Zetsma, CBSL, Tergooi MC, Department of Medical Microbiology, Hilversum

S. de Jager, Comicro, Department of Medical Microbiology, Hoorn

M.A. Leversteijn-van Hall, Eurofins Clinical Diagnostics, Department of Medical Microbiology, Leiden

M.T. van der Beek, Leiden University Medical Center, Department of Medical Microbiology, Leiden

S.P. van Mens, Maastricht University Medical Centre, Department of Medical Microbiology, Infectious Diseases and Infection Prevention, Maastricht

E. Schaftenaar, St Antonius Hospital, Department of Medical Microbiology and Immunology, Nieuwegein/Utrecht

J.C. Rahamat-Langendoen, Radboud University Medical Center, Department of Medical Microbiology, Nijmegen

P.D.J. Sturm, Laurentius Hospital, Roermond

B.M.W. Diederen, Bravis Hospital, Department of Medical Microbiology, Roosendaal

L.G.M. Bode, Erasmus University Medical Center, Department of Medical Microbiology and Infectious Diseases, Rotterdam

D.S.Y. Ong, Franciscus Gasthuis and Vlietland, Department of Medical Microbiology and Infection Control, Rotterdam

M. van Rijn, Ikazia Hospital, Department of Medical Microbiology, Rotterdam

S. Dinant, Maasstad Hospital, Department of Medical Microbiology, Rotterdam

M. den Reijer, Star-SHL, Rotterdam

D.W. van Dam, Zuyderland Medical Centre, Department of Medical Microbiology and Infection Control, Sittard-Geleen

E.I.G.B. de Brauwer, Zuyderland Medical Centre, Department of Medical Microbiology and Infection Control, Sittard-Geleen

A.L.E. van Arkel, Microvida ZorgSaam, Department of Medical Microbiology, Terneuzen

J.J.J.M. Stohr, Microvida Elisabeth-Tweesteden Hospital, Department of Medical Microbiology, Tilburg

A.L.M. Vlek, Diakonessenhuis, Department of Medical Microbiology and Immunology, Utrecht

M. de Graaf, Saltro Diagnostic Centre, Department of Medical Microbiology, Utrecht

A. Troelstra, University Medical Center Utrecht, Department of Medical Microbiology, Utrecht

F.N.J. Frakking, University Medical Center Utrecht, Department of Medical Microbiology, Utrecht

K.B. Gast, Eurofins-PAMM, Department of Medical Microbiology, Veldhoven

H.R.A. Streefkerk, VieCuri Medical Center, Department of Medical Microbiology, Venlo

S.B. Debast, Isala Hospital, Laboratory of Medical Microbiology and Infectious Diseases, Zwolle

## References

1. Lowy FD. Staphylococcus aureus infections. New England journal of medicine. 1998;339(8):520–532.

2. Ghalehnoo ZR. Diseases caused by Staphylococcus aureus. Int J Med Health Res. 2018;4(11):65–67.

3. Belgian Antibiotic Policy Coordination Commission, (BAPCOC). Guide belge de traitement anti infectieux en pratique ambulatoire/Belgische gids voor anti-infectieuze behandeling in de ambulante praktijk. 2022; https://overlegorganen.gezondheid.belgie.be/nl/documenten/belgische-gids-voor-anti-infectieuze-behandeling-de-ambulante-praktijk-2022.

4. National Institute for Health and Care Excellence, (NICE). Impetigo: antimicrobial prescribing. NICE guideline [NG153]. 2020; https://www.nice.org.uk/guidance/ng153.

5. Nederlands Huisartsen Genootschap, (NHG). Bacteriële huidinfecties (M68). [Bacterial skin infections]. 2019; https://richtlijnen.nhg.org/standaarden/bacteriele-huidinfecties.

6. Deplano A, Hallin M, Bustos Sierra N et al. Persistence of the Staphylococcus aureus epidemic European fusidic acid-resistant impetigo clone (EEFIC) in Belgium. Journal of Antimicrobial Chemotherapy. 2023;78(8):2061–2065.

7. Rijnders M, Wolffs P, Hopstaken R et al. Spread of the epidemic European fusidic acid-resistant impetigo clone (EEFIC) in general practice patients in the south of The Netherlands. Journal of antimicrobial chemotherapy. 2012;67(5):1176–1180.

8. Tveten Y, Jenkins A, Kristiansen B-E. A fusidic acid-resistant clone of Staphylococcus aureus associated with impetigo bullosa is spreading in Norway. Journal of Antimicrobial Chemotherapy. 2002;50(6):873–876.

9. de Greeff S, Kolwijck E, Schoffelen A. NethMap 2024. Consumption of antimicrobial agents and antimicrobial resistance among medically important bacteria in the Netherlands in 2023.2024.

10. Simonsen GS, Urdahl AM. NORM/NORM-VET 2016-Usage of Antimicrobial Agents and Occurrence of Antimicrobial Resistance in Norway. 2017.

11. Duarte ASR, Pessoa J, Attauabi M et al. DANMAP 2023: Use of antimicrobial agents and occurrence of antimicrobial resistance in bacteria from food animals, food and humans in Denmark. 2024.

12. Simonsen GSB, Blix HS, Helgesen KO et al. NORM/NORM-VET 2023. Usage of Antimicrobial Agents and Occurrence of Antimicrobial Resistance in Norway. 2024.

13. Vendrik KE, Kuijper EJ, Dimmendaal M et al. An unusual outbreak in the Netherlands: community-onset impetigo caused by a meticillin-resistant Staphylococcus aureus with additional resistance to fusidic acid, June 2018 to January 2020. Eurosurveillance. 2022;27(49):2200245.

14. Altorf-van der Kuil W, Schoffelen AF, de Greeff SC et al. National laboratory-based surveillance system for antimicrobial resistance: a successful tool to support the control of antimicrobial resistance in the Netherlands. Eurosurveillance. 2017;22(46):17–00062.

15. Schouls LM, Witteveen S, van Santen-Verheuvel M et al. Molecular characterization of MRSA collected during national surveillance between 2008 and 2019 in the Netherlands. Communications medicine. 2023;3(1):123.

16. Wertheim H, Vos M, Boelens H et al. Low prevalence of methicillin-resistant Staphylococcus aureus (MRSA) at hospital admission in the Netherlands: the value of search and destroy and restrictive antibiotic use. Journal of Hospital Infection. 2004;56(4):321–325.

17. Weterings V, Veenemans J, van Rijen M et al. Prevalence of nasal carriage of methicillin-resistant Staphylococcus aureus in patients at hospital admission in The Netherlands, 2010– 2017: an observational study. Clinical Microbiology and Infection. 2019;25(11):1428. e1421–1428. e1425.

18. Haute Autorité de Santé. Prise en charge des infections cutanées bactériennes courantes; Recommandation de bonne pratique. In: santé SdpidlfSfddHAd ed 2019.

19. Oranje AP, Chosidow O, Sacchidanand S et al. Topical retapamulin ointment, 1%, versus sodium fusidate ointment, 2%, for impetigo: a randomized, observer-blinded, noninferiority study. Dermatology. 2007;215(4):331–340.

20. Primhak S, Gataua A, Purvis D et al. Treatment of Impetigo with Antiseptics—replacing Antibiotics (TIARA) trial: a single blind randomised controlled trial in school health clinics within socioeconomically disadvantaged communities in New Zealand. Trials. 2022;23(1):108.

21. Nederlands Huisartsen Genootschap, (NHG). Toename van fusidinezuurresistente S. aureus. . 2024. https://www.nhg.org/actueel/toename-van-fusidinezuurresistente-s-aureus/.

