## Supplemental File for "Increasing trend in fusidic acid resistance among MRSA isolates in the Netherlands, 2016-2023"

### Supplementary material

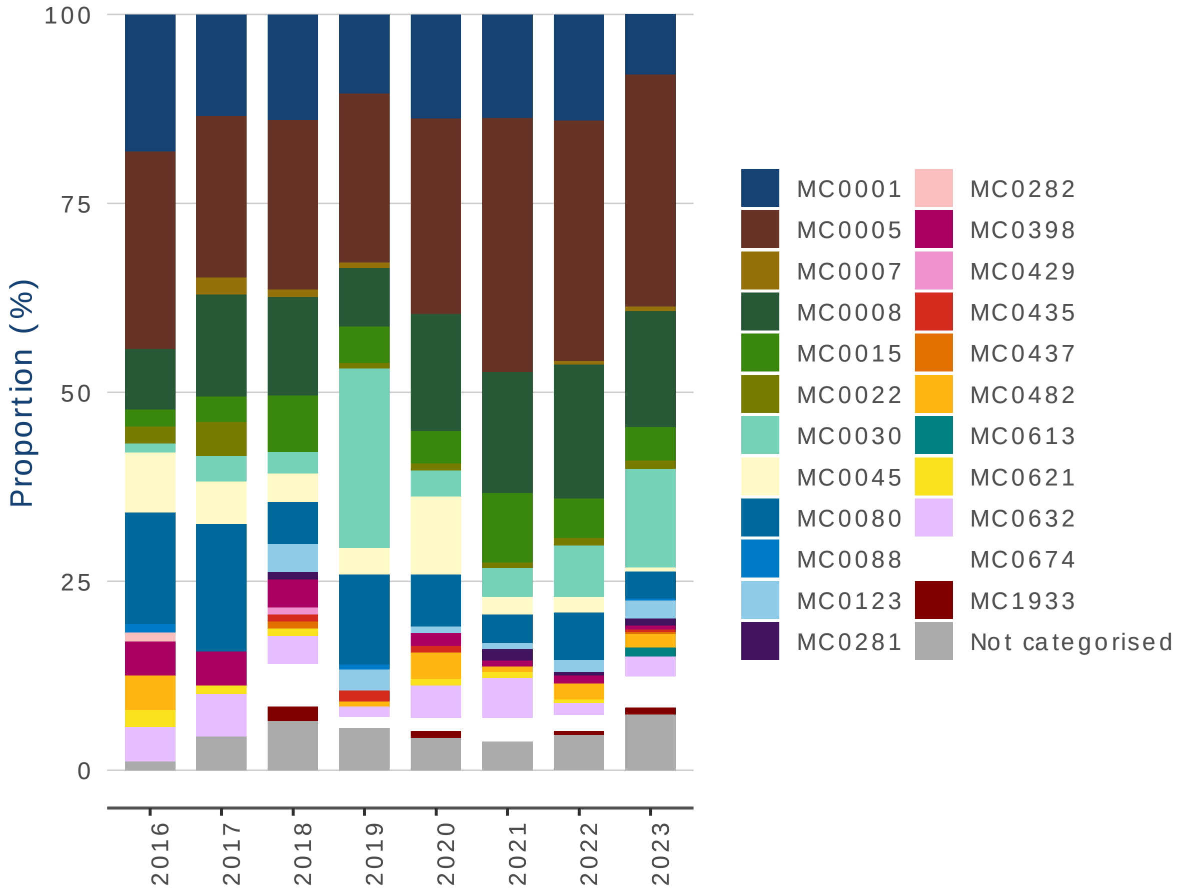


***Supplemental Figure 1 Proportion of MLVA-complexes among fusidic acid-resistant MRSA isolates between 2016 and 2023 in the Netherlands based on molecular typing data from Type-Ned.***

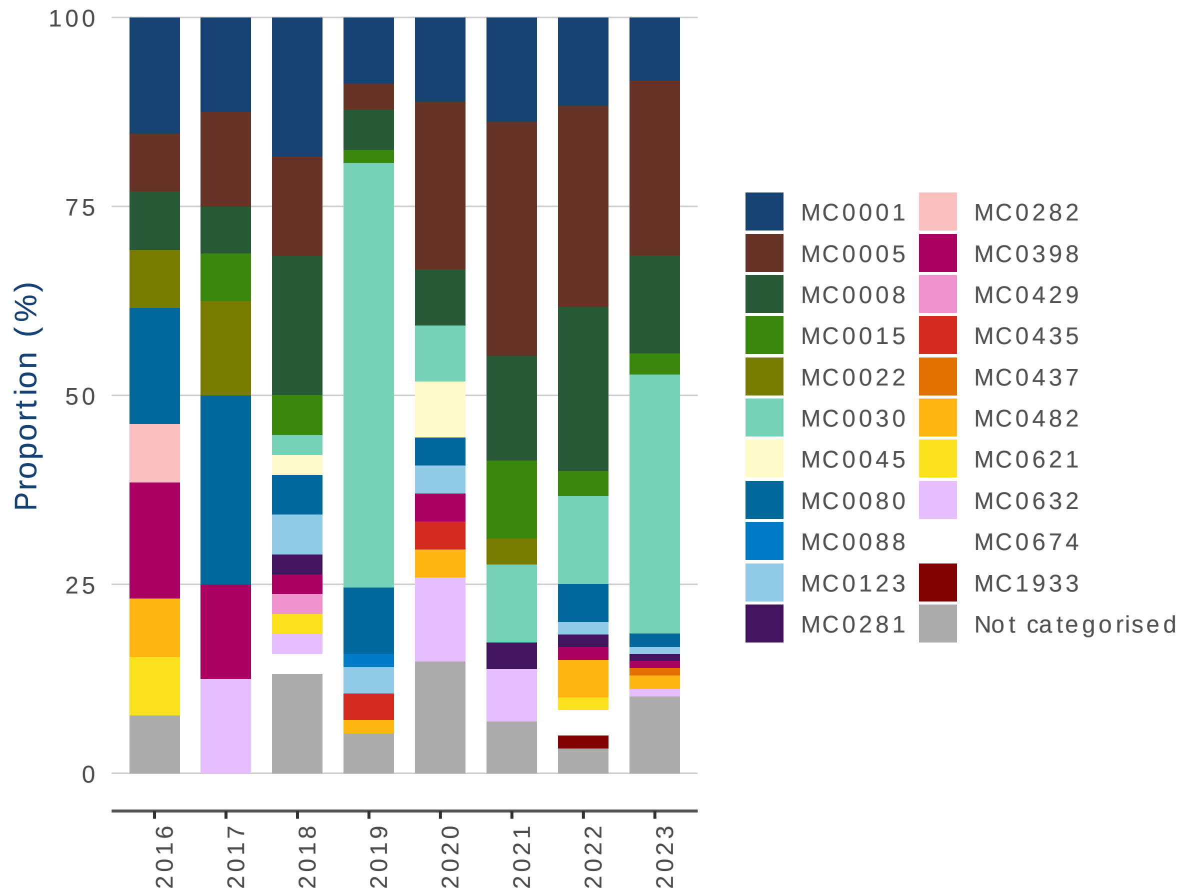


*Supplemental Figure 2 Proportion of MLVA-complexes among fusidic acid-resistant MRSA isolates from wound/pus/skin samples collected by the general practitioner between 2016 and 2023 in the Netherlands based on molecular typing data from Type-Ned.*

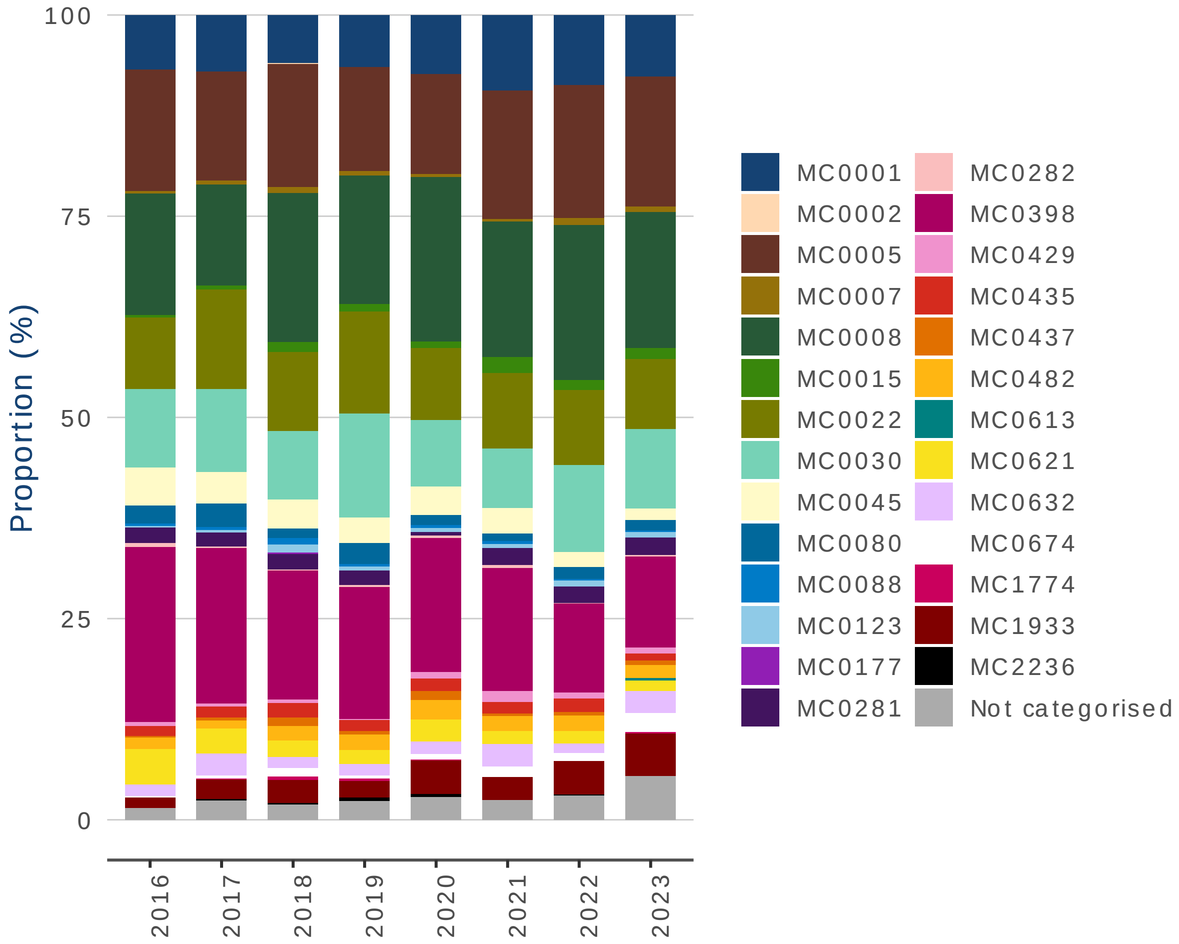


*Supplemental Figure 3 Proportion of MLVA-complexes among MRSA isolates between 2016 and 2023 in the Netherlands based on molecular typing data from Type-Ned.*

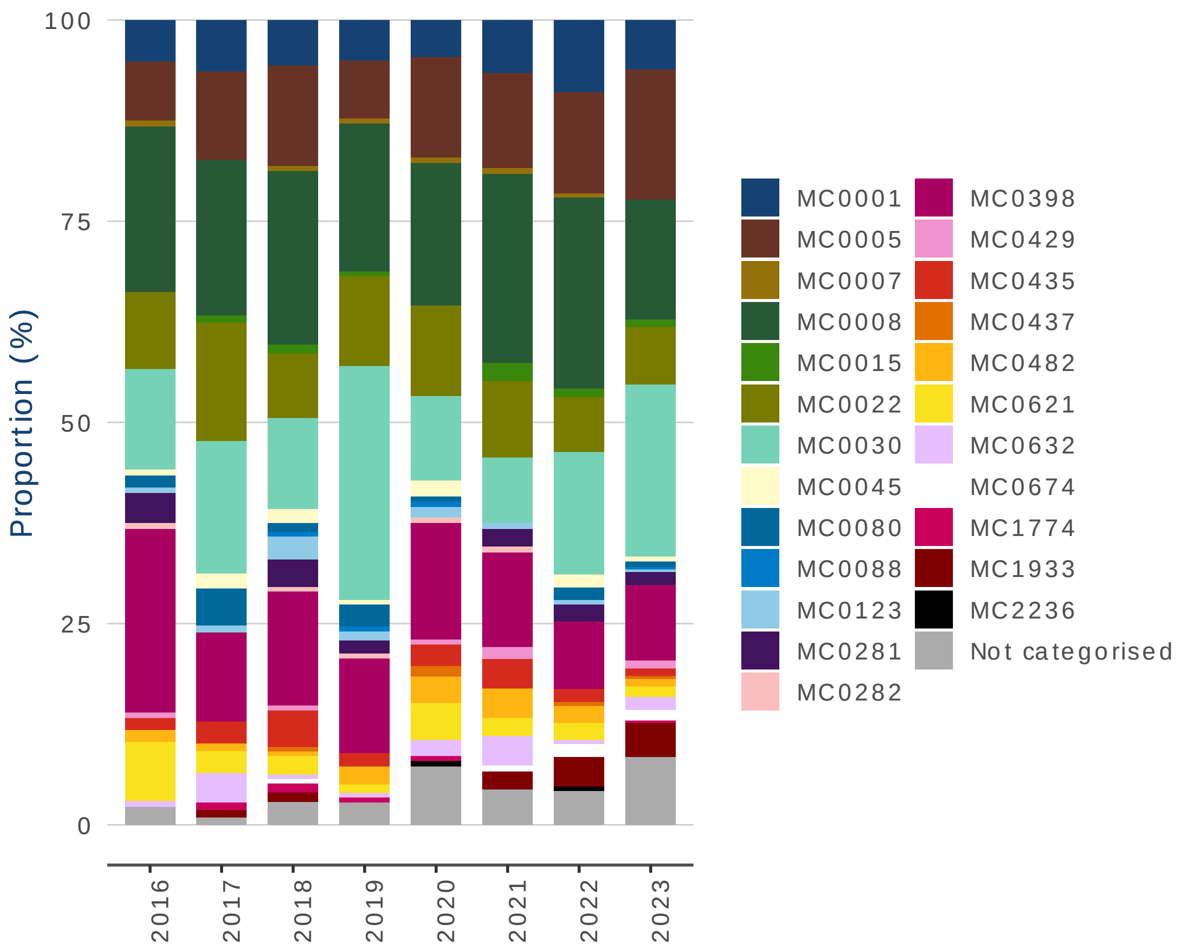


***Supplemental Figure 4 Proportion of MLVA-complexes among MRSA isolates from wound/pus/skin samples collected by the general practitioner between 2016 and 2023 in the Netherlands based on molecular typing data from Type-Ned.***

**Supplemental Table 1.** Odds ratios (ORs) from linear trends on fusidic acid resistance calculated per two year for MSSA and MRSA isolates overall and among WPS-GP samples.

| | MSSA – all isolates | MRSA – all isolates | MRSA – WPS-GP isolates | MSSA – WPS-GP isolates |
|---|---|---|---|---|
| **2018-2019 vs 2016-2017** | 1.08 (1.05-1.12)*** | 1.15 (0.96-1.38) | 2.09 (1.41-3.16)*** | 1.26 (1.18-1.35)*** |
| **2020-2021 vs 2016-2017** | 1.01 (0.98-1.05) | 1.30 (1.08-1.57)** | 1.52 (1.00-2.37) | 1.08 (1.01-1.16)* |
| **2022-2023 vs 2016-2017** | 1.10 (1.06-1.13)*** | 1.95 (1.66-2.31)*** | 3.12 (2.15-4.63)*** | 1.22 (1.15-1.30)*** |

*p<0.05
**p<0.01
***p<0.001
